# Variation across population subgroups of COVID-19 antibody testing performance

**DOI:** 10.1101/2020.09.14.20191833

**Authors:** Halley L. Brantley, Richard M. Yoo, Glen I. Jones, Marel A. Stock, Peter J. Park, Natalie E. Sheils, Isaac S. Kohane

**Affiliations:** UnitedHealth Group, Minnetonka, MN, USA; Department of Biomedical Informatics, Harvard Medical School, Boston, MA, USA

## Abstract

Understanding variations in the performance of serological tests for SARS-CoV-2 across varying demographics is relevant to clinical interpretations and public policy derived from their results. Appropriate use of serological assays to detect anti-SARS-CoV-2 antibodies requires estimation of their accuracy over large populations and an understanding of the variance in performance over time and across demographic groups. In this manuscript we focus on anti-SARS-CoV-2 IgG, IgA, and IgM antibody tests approved under emergency use authorizations and determine the recall of the serological tests compared to RT-PCR tests by Logical Observation Identifiers Names and Codes (LOINCs). Variability in test performance was further examined over time and by demographics. The recall of the most common IgG assay (LOINC 94563-4) was 91.2% (95% CI: 90.5%, 91.9%). IgA (LOINC 94562-6) and IgM (94564-2) assays performed significantly worse than IgG assays with estimated recall rates of 20.6% and 27.3%, respectively. A statistically significant difference in recall (*p* = 0.019) was observed across sex with a higher recall in males than females, 92.1% and 90.4%, respectively. Recall also differed significantly by age group, with higher recall in those over 45 compared to those under 45, 92.9% and 88.0%, respectively (*p*< 0.001). While race was unavailable for the majority of the individuals, a significant difference was observed between recall in White individuals and Black individuals (*p* = 0.007) and White individuals and Hispanic individuals (*p* = 0.001). The estimates of recall were 89.3%, 95.9%, and 94.2% for White, Black, and Hispanic individuals respectively.

## 1 Introduction

As the Coronavirus Disease 2019 (COVID-19) pandemic evolves, it is clear that testing is crucial for understanding the disease. Estimates of disease prevalence can be used to inform public policy and help prioritize scientific research agendas, but accurate estimates require the correct interpretation of testing data. They also directly affect individual patients to the extent that clinicians assume negative test results are equivalent to no viral exposure. If systematic differences in test performance across different population subgroups exist, they should, at the very least, inform both policy and clinical decision-making.

We analyze here a collection of almost 2.5 million test results to assess the quality of antibody tests (see Table 1). Specifically, a positive SARS-CoV-2 reverse transcription polymerase chain reaction (RT-PCR) test is assumed to be the reference gold standard for COVID-19 infection, to which antibody test results are compared against. Given the limited supporting evidence on these tests and their variable performance depending on the stage of the disease, this approach provides a simplified framework for determining the recall of serological antibody tests [1,2].

**Table 1.** Dataset summary.

|  |  |
| --- | --- |
| Date range for testing | March 2, 2020 through August 1, 2020 |
| Number of individuals tested | 1,913,640 |
| Age range | 1 month to >100 years old |
| Gender | 56.1% female, 43.6% male |
| Total number of tests | 2,445,907 |
| Test type given | 66.8% PCR tests, 33.2% antibody tests |

## 2 Methods

### 2.1 Data

We used de-identified administrative claims for individuals enrolled in a Medicare Advantage, commercial, or Medicaid plan in a research database from a single large health insurance provider in the United States. The database contains medical (physician, inpatient, outpatient) and pharmacy claims for services submitted for third-party reimbursement. We joined this claims database with a limited outpatient dataset that included test results for individuals undergoing outpatient testing for SARS-CoV-2 at 84 hospital-based, free-standing outpatient, and third-party labs across the United States. Approximately 90% of the test results were submitted by just two large national labs (Lab A: 58.5% and Lab B: 31.0%), with other 82 labs making up the final 10.5% of tests. Results for 1,864 individuals were excluded from the analysis due to multiple reported sexes or birth dates. The resulting dataset included 2,445,907 test results from 1,913,640 individuals (Table 1). The population represents a wide range of ages, and a good balance of sex. Figure 1 presents the count of individuals by state, showing that the dataset covers individuals from across the United States with the highest number of tests conducted in New York, followed by Florida, and Texas.

**Figure 1.**
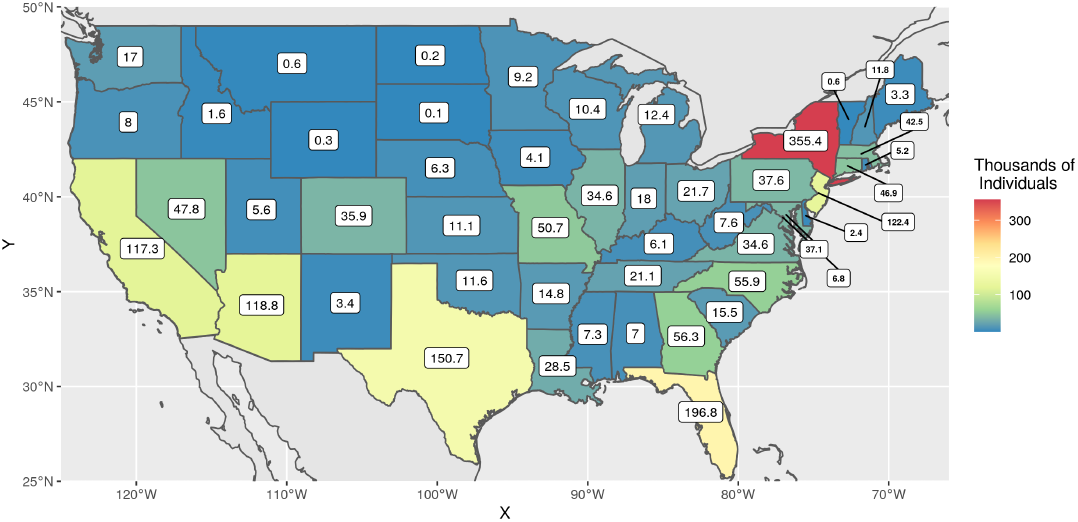
Counts of individuals tested by state in our dataset. Not shown: Alaska (*n* = 384), Hawaii (*n* = 1, 267), and Washington D.C. (*n* = 6, 755).

The tests themselves are identified by Logical Observation Identifiers Names and Codes (LOINCs). The LOINCs associated with the tested population and the frequency of tests reported are listed in Table 2. It is notable that 65.54% of all tests conducted were RT-PCR tests of one particular type (LOINC 94500-6). This LOINC accounted for 98.12% of RT-PCR tests and is associated with tests manufactured by over 40 different entities [3]. As LOINCs often exist in a one-to-many relationship to manufacturers, the data does not allow for analysis of specific tests marketed by a particular manufacturer. To get the most robust estimate of the recall of antibody tests, only RT-PCR tests with LOINC 94500-6 are considered in the following analysis.

**Table 2.** Administered tests by LOINC code.

| Type | Component | LOINC | Lab A | Lab B | Other | Total | All Tests | Same Type |
| --- | --- | --- | --- | --- | --- | --- | --- | --- |
| Serological | Ab | 94661-6 | 0 | 0 | 15 | 15 | 0.00% | 0.00% |
|  |  | 94762-2 | 0 | 11,389 | 41,668 | 53,057 | 2.17% | 6.53% |
|  | Ab IgA | 94562-6 | 0 | 20,371 | 225 | 20,596 | 0.84% | 2.54% |
|  | Ab IgG | 94505-5 | 0 | 0 | 2,435 | 2,435 | 0.10% | 0.30% |
|  |  | 94507-1 | 0 | 0 | 4,023 | 4,023 | 0.16% | 0.50% |
|  |  | 94563-4 | 420,435 | 217,762 | 36,081 | 674,278 | 27.57% | 83.01% |
|  | Ab IgG+IgM | 94547-7 | 0 | 0 | 875 | 875 | 0.04% | 0.11% |
|  | Ab IgM | 94508-9 | 0 | 0 | 593 | 593 | 0.02% | 0.07% |
|  |  | 94564-2 | 0 | 55,506 | 907 | 56,413 | 2.31% | 6.94% |
|  | Total | Total | 420,435 | 305,028 | 86,822 | 812,285 | 33.21% | 100.00% |
| PCR | N gene | 94316-7 | 0 | 0 | 1,509 | 1,509 | 0.06% | 0.09% |
|  |  | 94533-7 | 0 | 0 | 684 | 684 | 0.03% | 0.04% |
|  | RdRp gene | 94534-5 | 0 | 0 | 142 | 142 | 0.01% | 0.01% |
|  | RNA | 94309-2 | 7,556 | 0 | 20,028 | 27,584 | 1.13% | 1.69% |
|  |  | 94500-6 | 1,003,562 | 454,062 | 145,326 | 1,602,950 | 65.54% | 98.12% |
|  |  | 94559-2 | 0 | 0 | 496 | 496 | 0.02% | 0.03% |
|  |  | 94565-9 | 0 | 0 | 184 | 184 | 0.01% | 0.01% |
|  |  | 94759-8 | 0 | 0 | 73 | 73 | 0.00% | 0.00% |
|  | Total | Total | 1,011,118 | 454,062 | 168,442 | 1,633,622 | 66.79% | 100.00% |
|  | Grand Total | Grand Total | 1,431,553 | 759,090 | 255,264 | 2,445,907 | - | - |

Of the 130,835 individuals with positive RT-PCR tests, 191 individuals had a negative RT-PCR test result reported on the same day as their positive result and were removed from the analysis; this left 130,644 individuals with a valid positive RT-PCR test. The positive RT-PCR tests occurred throughout the date range (March 2, 2020 through August 1, 2020) but are concentrated in April and July (Figure 2). Of note, we do not have access to testing results for at home and point-of-care lateral flow assay tests.

**Figure 2.**
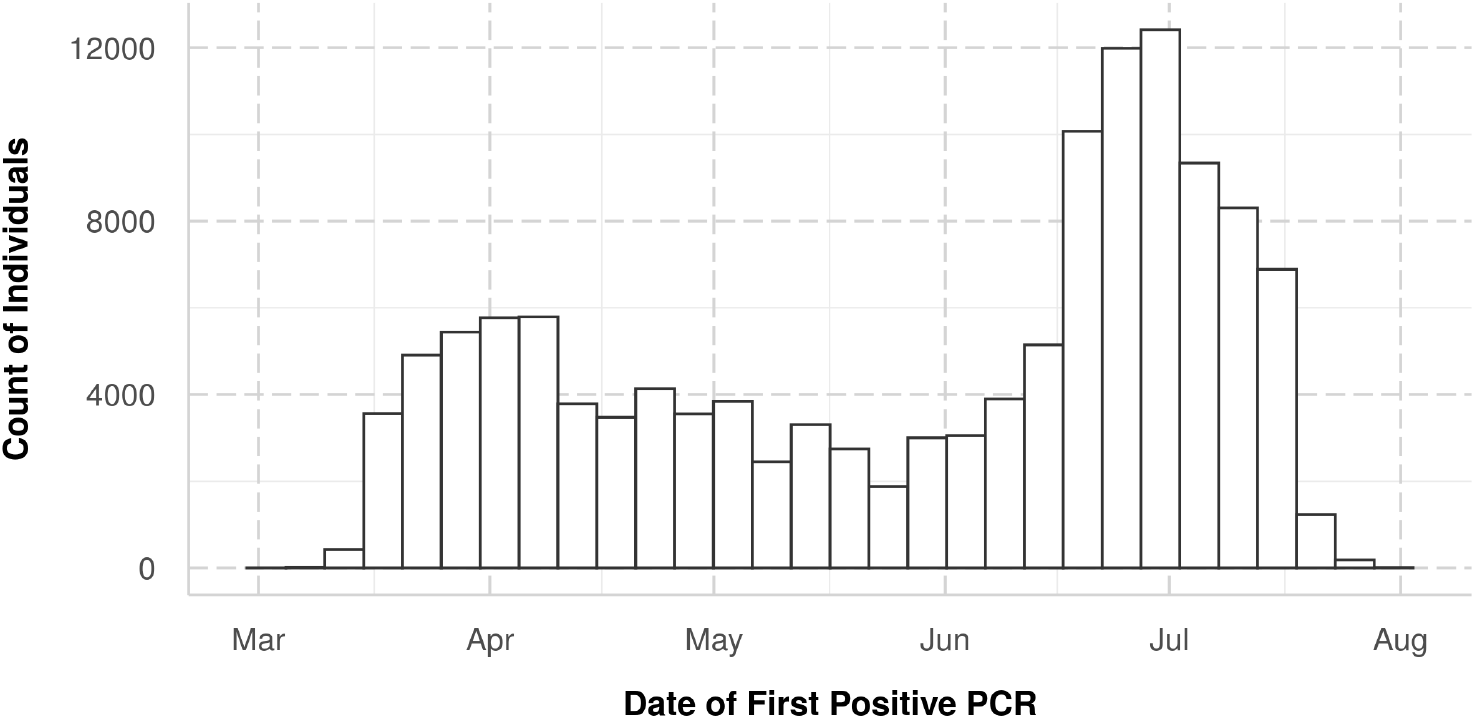
Counts of individuals with positive RT-PCR tests by date.

Like the RT-PCR tests, the antibody tests also concentrated into a single LOINC code with 83.01% of them coded as LOINC 94563-4 (Table 2). The next most frequent antibody test (LOINC 94564-2) only accounted for 6.94% of antibody tests. While there were still thousands of tests conducted using the 2nd through 6th ranked tests, they represented relatively small samples after filtering was applied.

### 2.2 Data Analysis

When estimating recall, only individuals that received a positive RT-PCR test and any antibody test were considered. When estimating recall overall and by demographics, antibody test results were only considered if they succeeded the first positive RT-PCR test by at least 14 days. This provides sufficient time for seroconversion to occur and be reasonably detectable by an antibody test as antibodies may take an average of 10 to 14 days to be detectable after infection [4–10]. Additionally, only antibody tests conducted on the first date that met the criteria were considered to avoid bias by counting a single individual multiple times. When estimating recall changes by prognosis, all antibody tests were considered.

Confidence intervals for recall were calculated by inverting the score test with the null hypothesis that the recall was equal to 0.5. Yates’ continuity correction was applied where possible and the resulting interval was clipped to [0, 100]. All analyses were conducted using the statistical software R [11]. Proportion tests were used to identify significant differences across sex, race, and age group. A total of eight hypothesis tests were conducted and the p-values were adjusted using the Benjamini-Hochberg method to control the false discovery rate [12].

While recall estimates were calculated for all antibody LOINCs with more than 10 individuals with positive results, we focused demographic analyses on individuals with the most frequent antibody test (LOINC 94563-4) to avoid confounding of the results by the use of less sensitive tests.

This study was reviewed and deemed exempt by the institutional review board of UnitedHealth Group.

## 3 Results

### 3.1 Antibody Testing Recall

Figure 3 shows population counts by test type and result. Filtering the data down to include only those individuals with a positive RT-PCR test at least 14 days prior to an antibody test and excluding an individual with conflicting antibody test results on the same date created a pool of 7,406 people. The demographics in this subset were similar to those in the complete dataset with a sex split of 47/53% (male/female) and an age range of 1 to over 100 years old with a median age of 52. The population represents 48 states with the highest counts in New York, New Jersey, and Florida.

**Figure 3.**
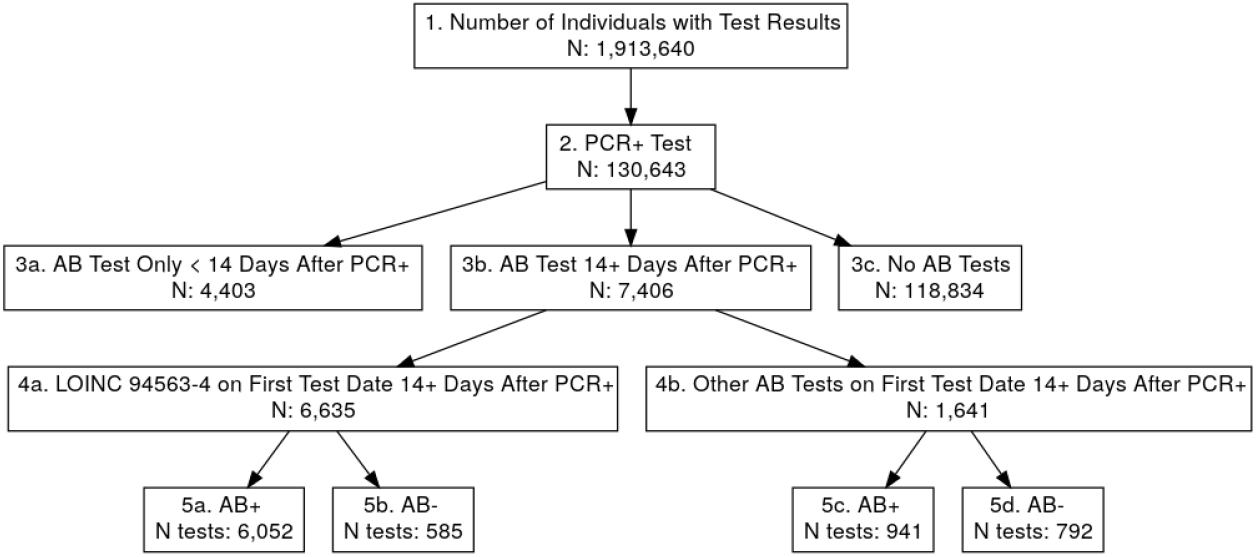
Counts of distinct individuals by type of test and results. Counts in row 5 refer to number of tests rather than individuals. One person had conflicting LOINC 94563-4 test results and was removed from the population. Boxes 4a and 4b do not sum to 3b because 871 individuals received both a LOINC 94563-4 test result and another antibody (AB) test result on the first testing date that met the criteria.

The time from the positive RT-PCR test to the first antibody test ranged from 14 to 133 days with a median of 38 days and mean of 43 days. A total of 871 individuals received both a LOINC 94563-4 test result and another antibody test result on the first testing date that met the criteria. Of the remaining pool, 6,635 individuals received an IgG antibody test recorded as LOINC 94563-4, and 6,052 of those results were positive, resulting in a recall of 91.2% and 95% confidence interval of (90.5%, 91.9%).

Despite the relatively high recall rate of LOINC 94563-4, performance was highly variable across the nine types of serologic tests used^1^. Table 3 reports recall rates for these tests and their 95% confidence intervals. Estimates of average test recall rates range from a low of 20.6% (LOINC 94562-6) to a high of 91.2% (LOINC 94563-4). The poorest-performing LOINC 94562-6 measures IgA antibodies. IgA is primarily secreted from mucosal membranes, where it fights early viral entry into cells [13]. There has been speculation that testing for the IgA antibody may not be a reliable method for identifying COVID-19 infections, and these findings seem to validate this as it is the poorest-performing test. The test that measures IgM also performs significantly worse than the IgG tests with a recall of 27.3% for LOINC 94564-2.

**Table 3.** Estimated recall of serological tests by LOINC. Only LOINCs with > 10 positive results reported are shown.

| LOINC | Component | Total Count | Positive Count | Recall | Lower Bound | Upper Bound |
| --- | --- | --- | --- | --- | --- | --- |
| 94563-4 | Ab IgG | 6,635 | 6,052 | 91.2% | 90.5% | 91.9% |
| 94564-2 | Ab IgM | 904 | 247 | 27.3% | 24.5% | 30.4% |
| 94762-2 | Ab | 706 | 638 | 90.4% | 87.9% | 92.4% |
| 94562-6 | Ab IgA | 296 | 61 | 20.6% | 16.2% | 25.8% |
| 94507-1 | Ab IgG | 15 | 13 | 86.7% | 58.4% | 97.7% |

### 3.2 Recall Changes Over Prognosis

Relaxing the 14 day requirement for antibody tests resulted in a population of 11,810 individuals with 15,232 reported test results. From what is currently understood about the timing of serological conversion, we expect that the probability of a positive antibody test would increase over time followed by a decrease as antibody titers decline. In general, the data supported this hypothesis (Figure 4), with most tests exhibiting the highest recall rates after three and four weeks from the date of the positive RT-PCR test.

**Figure 4.**
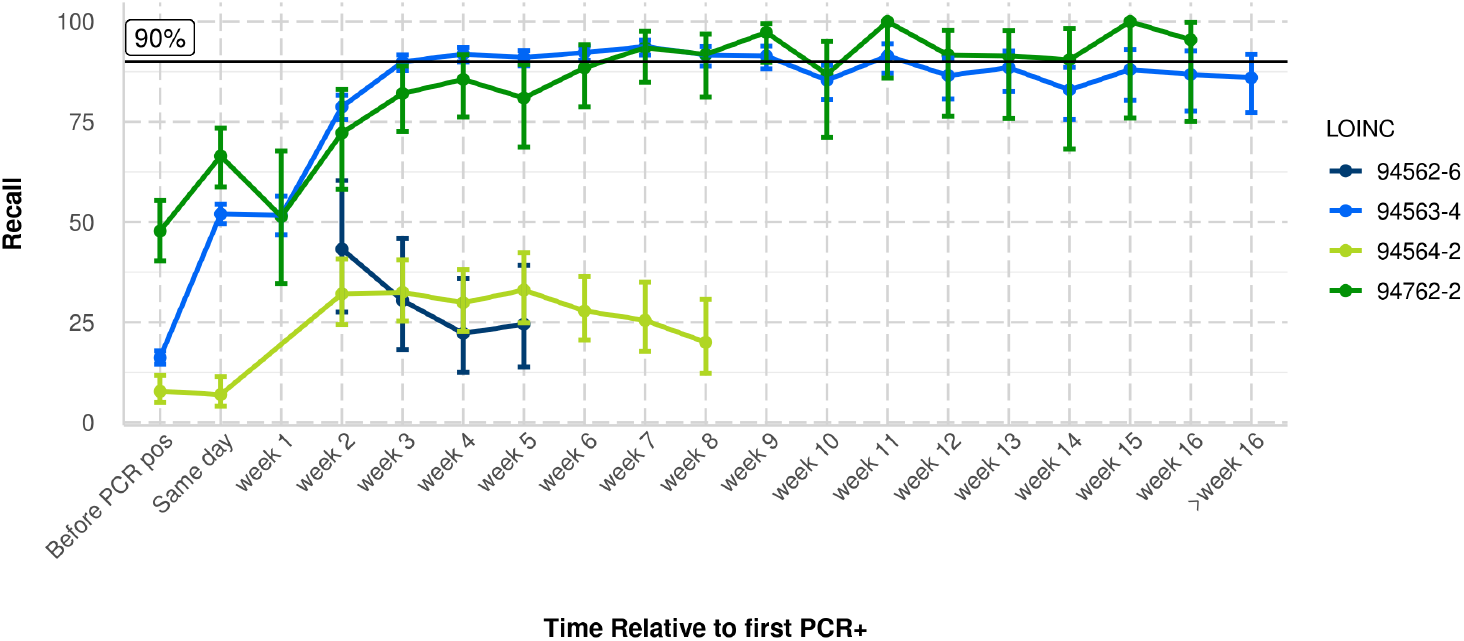
Estimated recall of serological tests by length of time from first positive RT-PCR test. Only groupings with > 10 positive results reported are shown.

However, the degree to which recall diminishes after that peak varied greatly across tests. The most common IgG test (LOINC 94563-4) maintains recall above 90% from week three to week nine peaking at week seven with samples sizes ranging from 91 to 964 for weeks one through 16. Only in week ten and week 14 does the 95% confidence interval fall below 90%. The estimated recall of the non-specific antibody test (LOINC 94762-2) rises above 90% at week 7 and the upper bound of the confidence interval stays above 90% through week 16. The sample sizes for LOINC 94762-2 are smaller ranging from 16 to 95 for weeks one through 16. The recall of the IgA test (LOINC 94562-6) decreases dramatically from week two to week five, while the recall of the IgM test (LOINC 94564-2) steadily decreases from week five to week eight.

As the starting time of this analysis is RT-PCR testing, the variability of the intervals noted above depends where in the course of infection these first tests occurred. For example late RT-PCR testing might detect viral fragments even after live virus is cleared and the subsequent IgG assay may appear positive early relative to others. Interestingly, for the most common test LOINC 94563-4 (IgG), 1,993 antibody tests were conducted prior to a RT-PCR test, with 16.1% of the results being positive. When LOINC 94563-4 was taken on the same day as the positive RT-PCR, 52% of the results were positive.

### 3.3 Recall Across Demographics

The demographic analysis was restricted to the most common IgG test (LOINC 94563-4). Table 4 shows the breakdown of recall rate across sex, age group, race, and income group.

Breakdown by sex showed a statistical difference in the implied recall rates (*p* = 0.019), where the male population exhibited higher recall.

**Table 4.** Estimated recall of serological tests by demographics. Complete demographic information was not available for all individuals, only non-missing data is displayed.

|  |  | Total Count | Positive Count | Recall | Lower Bound | Upper Bound |
| --- | --- | --- | --- | --- | --- | --- |
| Gender | female | 3,526 | 3,186 | 90.4% | 89.3% | 91.3% |
|  | male | 3,105 | 2,861 | 92.1% | 91.1% | 93.1% |
| Age Group | [0,12) | 28 | 26 | 92.9% | 75.0% | 98.8% |
|  | [12,25) | 435 | 373 | 85.7% | 82.0% | 88.8% |
|  | [25,35) | 814 | 707 | 86.9% | 84.3% | 89.1% |
|  | [35,45) | 1,034 | 927 | 89.7% | 87.6% | 91.4% |
|  | [45,55) | 1,365 | 1,263 | 92.5% | 91.0% | 93.8% |
|  | [55,65) | 1,569 | 1,461 | 93.1% | 91.7% | 94.3% |
|  | [65,75) | 960 | 896 | 93.3% | 91.5% | 94.8% |
|  | [75,85) | 331 | 305 | 92.1% | 88.6% | 94.7% |
|  | [85,105] | 99 | 93 | 93.9% | 86.8% | 97.5% |
| Race | White | 980 | 875 | 89.3% | 87.1% | 91.1% |
|  | Hispanic | 800 | 754 | 94.2% | 92.3% | 95.7% |
|  | Black | 242 | 232 | 95.9% | 92.3% | 97.9% |
|  | Asian | 53 | 51 | 96.2% | 85.9% | 99.3% |
|  | Other | 172 | 160 | 93.0% | 87.9% | 96.2% |
| Income Group | <\$50K | 1,194 | 1,099 | 92.0% | 90.3% | 93.5% |
| | \$50-\$75K | 1,977 | 1,791 | 90.6% | 89.2% | 91.8% |
| | \$75 - \$100K | 1,555 | 1,420 | 91.3% | 89.8% | 92.6% |
| | >\$100K | 1,774 | 1,618 | 91.2% | 89.8% | 92.5% |

Breakdown of recall rate across age group showed increased recall for elder groups, with ages 12 to 25 and 25 to 35 exhibiting the lowest recall rates, 85.7% and 86.9%, respectively^2^.

Recall was also broken down by age and sex simultaneously (Figure 5). The increased recall in males persists across all age groups except the youngest and eldest for which sample sizes are the smallest. Recall appears to increase with age until around age 45. A statistically significant difference was observed between the recall in those 45 and above, 92.9%, compared with those under 45, 88.0% (*p*< 0.001).

**Figure 5.**
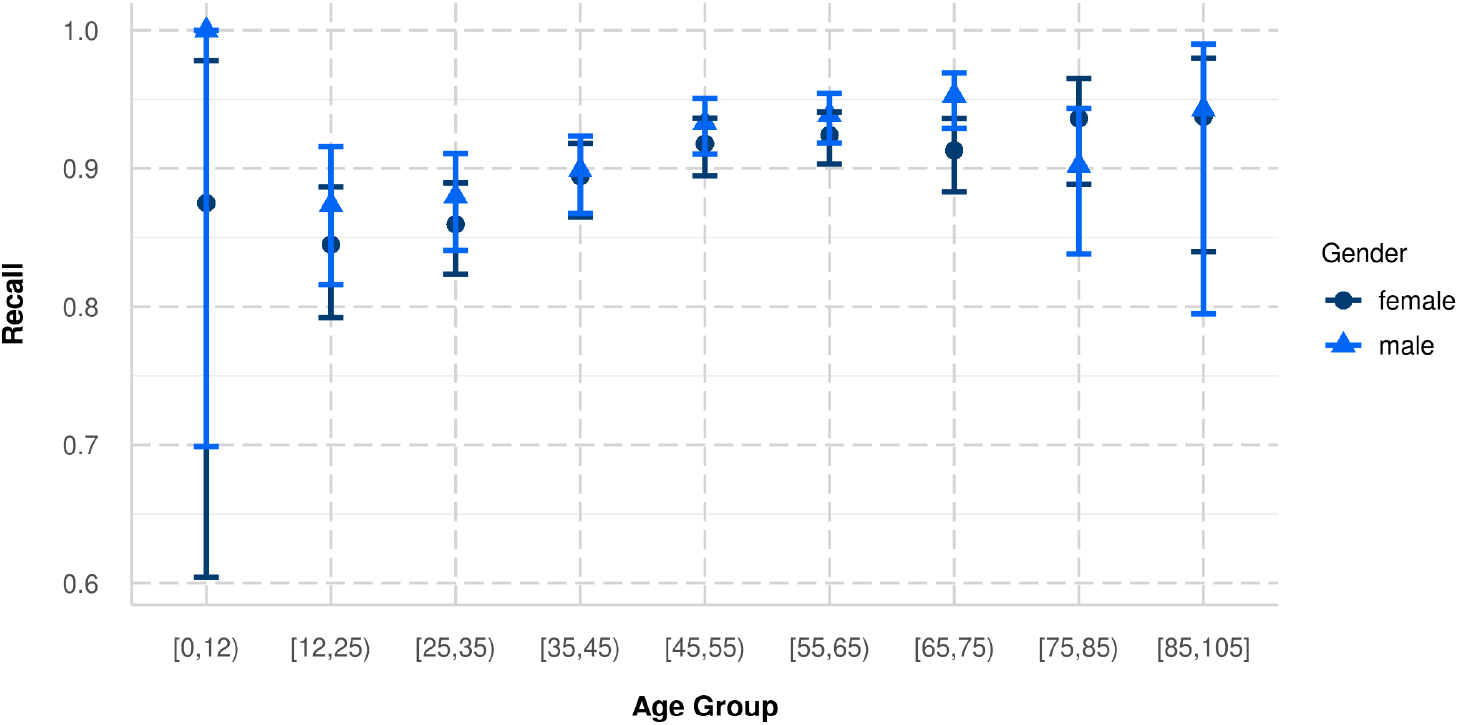
Estimated recall of serological tests by age and sex.

While race was unknown for the majority of individuals, available data suggests a lower recall in White individuals than in Hispanic or Black individuals; between White individuals and Black individuals (*p* = 0.007) and White individuals and Hispanic individuals (*p* = 0.001). The significant differences by race persist when the population is restricted to those aged 45 and above.

Income group was assigned based on the most recent median household income in the individual’s zipcode, using the method described in [14]. Recall did not vary significantly by income group.

## 4 Discussion

While the FDA has issued emergency use authorization [15] for many tests during this pandemic, the amount of data supporting these authorizations is frequently limited. This is especially true of serological tests for antibodies; most recent analysis results [10,16,17] are over tens of individuals. A study that quantifies their accuracy over a large population has been lacking.

Our study found important differences between IgG and IgM tests in contrast to no apparent differences found in the meta-analysis performed in [8]. Of note, this meta analysis included much smaller sample count (5,016 samples) compared to the 2.4 million samples in our study, but also the time intervals encompassed by our studies differ. Findings in [9] also support our conclusion that tests measuring IgM are less reliable than those measuring IgG.

Overall, the empirically derived IgG antibody test recall (LOINC 94563-4) is 91.2%, assuming a positive RT-PCR test as the gold standard. IgM and IgA tests performed significantly worse than IgG and non-specific tests. There is additional variability in recall, likely related to the variation in peak antibody levels in individuals, with respect to sex, age, and ethnicity. We found a recall difference across sex (*p* = 0.019) and racial groups; between White individuals and Black individuals (0.007), and White individuals and Hispanic individuals (*p* = 0.001) but no significant difference based on income as estimated from zipcode. Recall increased steadily and significantly with age until 45 years of age, with a significant difference observed between those 45 and above compared with those under 45 (*p*< 0.001).

Our results are similar to prior studies detecting viral presence of SARS-CoV-2. For example, Gudbjartsson et al. [18] found that children under 10 years old were less likely to test positive as were females. These differences could be due to different ascertainment or referral patterns and timing of the ascertainment in these subpopulations. The well documented differences of prevalence in these populations [19,20] do not appear to explain the differences in test performance. Alternative, biological hypotheses for these differences in seropositivity recall with control for confounding by co-existing conditions such as obesity [21,22], which have been shown to be significantly associated with reduced antibody responses, will be pursued. Evidence for these biological hypotheses (e.g. Takahashi et al. [23–26]) is very preliminary and requires specific validation for human SARS-CoV-2 infections.

### 4.1 Limitations

This study has many of the limitations of retrospective observational studies, not least of which is how the patients were ascertained. This outpatient testing data is skewed since individuals who receive these tests are generally healthier and have relatively fewer comorbid conditions than the general population. Biases are especially likely in the earliest stage of the pandemic when the supply of tests was limited and testing was frequently reserved for those deemed most likely to be infected. The high number of asymptomatic infections is another consideration for understanding the tested population, as those without symptoms may have been less likely to seek out or be given a test. In fact, some reports show a lower peak antibody level response in these patients [4]. Also, using the RT-PCR test as a gold standard means that this study does not include the serology of those patients who had a false negative RT-PCR test, which may be substantial given reports of poor sensitivity [27,28]. Nonetheless, the recall rates we report here do reflect much of current practice relative to positive RT-PCR assays, and the subpopulation seropositivity recall differences are therefore of interest to policy-making and clinical practice.

## Data Availability

The data are proprietary and are not available for public use but can be made available under a data use agreement to confirm the findings of the current study. The statistical code is available from the first author.

## 5 Acknowledgment

## A Database Quality

### A.1 Standardization of Data Entry and Data Structure

Medical and pharmacy claims data are captured, predominantly electronically, from sites of care seeking third-party reimbursement for both Medicare and commercial plans using the industry standard data collection forms HCFA/CMS-1500 for facility claims, UB04/CMS-1450 for professional services and outpatient claims, and NCPDP for pharmacy claims or their electronic equivalents. Structured data from these standardized forms are coded using the International Classification of Diseases, Tenth Revision, Clinical Modification (ICD-10-CM), National Drug Codes (NDC), Current Procedural Terminology (CPT) codes, and Logical Observation Identifiers Names and Codes (LOINC) codes, and Diagnosis Related Groups (DRG). This nomenclature ensures consistency of data collection across geographic regions, health systems, and payers throughout the United States.

### A.2 Methods to Control for Errors in Sampling and Data Collection

Claims that do not adhere to the form or coding standards described above are rejected from reimbursement, minimizing the risk that inappropriately structured data are included in the database. Data specific to SARS-CoV-2 and COVID-19 has an additional Quality Control layer to control for errors in sampling and data collection; this is described below in Section A.7.

### A.3 Data Relevance and Accuracy

Data are transferred into the UnitedHealth Group (UHG) Clinical Discovery Database, where a dedicated team pursues data management to ensure accurate matching of source data to an individual. This protocol uses unique identifiers to match them to existing identifiers in the UHG Clinical Discovery Database to determine whether the individual already exists in the platform. A unique identification number is generated for each individual so that data from multiple sources can be linked back to that identification number. Individuals that fail to meet the matching criteria are excluded from the UHG Clinical Discovery Database to reduce the risk of erroneous linkage of records. Those whose claims do not fulfill basic standardized data structure requirements described previously are also excluded. During this, all member protected data are stored in a separate database that is only accessible by a designated engineering team. In addition to a persistent identifier being generated for each member, a de-identified primary key is also generated. The de-identified primary key is recycled every 6 months, at which time each member is assigned a new de-identified primary key. Data that are made available for research through the UHG Clinical Discovery Database use the de-identified primary key as the link across data tables. All protected information has been removed, ensuring any research performed is limited to retrospective analysis of de-identified data and accessed in accordance with Health Insurance Portability and Accountability Act regulations.

### A.4 Sufficiency of Basic Data

As described above, individuals lacking enough data to be assigned a unique primary key are excluded from the UHG Clinical Discovery Database, as are patients whose claims did not fulfill basic data structure requirements. In a given month in 2019, the UHG Clinical Discovery Database contained one or more claims from 5 million Medicare Advantage enrollees and 20 million commercially-insured individuals. Further information on data sufficiency for the research performed in this manuscript can be found in Figure 3.

### A.5 Adequacy of Possible Derived Data

To reduce the risk of introducing error to standardized, structured claims data, derivation of source data within the UHG Clinical Discovery Database is minimal. The Data Integration team loads, formats, and join the data to appropriate dimension tables. Dimension tables are combined with raw claims information to limit the number of times external tables need to be referenced. Researchers may request derived fields within data tables prepared specifically for a project. This process is managed by the Data Enrichment team, who creates data dictionaries to accompany derived fields. Tables containing derived data are stored separately from raw source data.

### A.6 Design of Computer Editing Methods

Access to modify/edit source data is restricted to a subset of data specialists. Each step in the data flow has a restricted list of individuals able to perform any type of editing to the database, and access level varies by team (Data Integration, Data Enrichment). Researchers using the Clinical Discovery Database may not edit any source data or enrichment data. They are instead given access to “sandbox” locations where they may request editing access for the data tables used in their analyses.

### A.7 Quality Control

In addition to the quality control mechanisms described during the matching procedures to reject non-linkable or inappropriately structured data, a COVID-19 data source-specific layer of quality control is also present, given the rapidly evolving situation. SARS-CoV-2 lab tests included in the UHG Clinical Discovery Database exclude custom local codes or codes that are not present in the LOINC organization’s guidance for mapping SARS-CoV-2 and COVID-19 related LOINC terms. Test information provided via the LOINC code compliments the test type (antibody, RT-PCR, etċ as well as the result value (detected, not detected, not given/cancelled). Suspected COVID-19 inpatient cases are manually reviewed daily by health plan clinical staff via clinical notes to determine an individual’s COVID-19 status. Each case is then manually flagged as either negative, confirmed, presumed positive, or needs clinical review. If a case is confirmed, it is not reviewed again. If a case is listed as negative or unknown, it is periodically reviewed for changes in the record. All others are reviewed and updated daily.

### A.8 Differences Across Groups

While the data for Medicare Advantage and commercially insured enrollees is processed in a similar manner, these groups are substantially different. First, there are systematic differences in patient characteristics, most remarkably the older age and the higher prevalence of all comorbidities. These differences are tabulated in the UHG Clinical Discovery Database, there are restrictions from individual employers on these use of data for research. Therefore, commercial insurance claims that are available for analyses are a subset of the overall commercially insured population.

### A.9 Data Sharing

The data are proprietary and are not available for public use but can be made available to editors and their approved auditors under a data use agreement to confirm the findings of the current study.

## Footnotes

1 Four results were removed from the analysis due to small sample size (≤ 10 positive results).

2 High recall and the ensuing variability for the youngest group is due to sample size.

